# How early into the outbreak can surveillance of SARS-CoV-2 in wastewater tell us?

**DOI:** 10.1101/2020.08.19.20177667

**Authors:** Pei-Ying Hong, Andri Taruna Rachmadi, David Mantilla-Calderon, Mohsen Alkahtani, Yasir M. Bashwari, Hamed Al Qarni, Jianqiang Zhou

**Affiliations:** Division of Biological and Environmental Science and Engineering, Water Desalination and Reuse Center, King Abdullah University of Science and Technology (KAUST), Thuwal 23955–6900, Saudi Arabia; Environmental Health Laboratory, Jeddah, Ministry of Health, Saudi Arabia; General Directorate of Environment Health, Ministry of Health, Saudi Arabia

**Keywords:** Wastewater-based epidemiology, Covid-19, reverse transcription quantitative PCR, detection sensitivity, log reduction

## Abstract

There is increasing interest to use wastewater-based surveillance of SARS-CoV-2 as an early warning of the outbreak within a community. Despite successful detection of SARS-CoV-2 in wastewaters sampled from multiple locations, there is still no clear idea on the minimal number of cases needed in a community to result in a positive detection of the virus in wastewaters. To address this knowledge gap, we sampled wastewaters from a septic tank and biological activated sludge tank located on-site of a hospital. The hospital is providing treatment for SARS-CoV-2 infected patients, with the number of hospitalized patients per day known. It was observed that > 253 positive cases out of 10,000 persons are required prior to detecting SARS-CoV-2 in wastewater. There was a weak correlation between N1 and N2 gene abundances in wastewater with the number of hospitalized cases. This correlation was however not observed for N3 gene. The occurrence frequency of SARS-CoV-2 is at least 5 times lower in the partially treated wastewater than in the septic tank. Furthermore, abundance of N1 and N3 genes in the activated sludge tank were 50 and 70% of the levels detected in septic tank, suggesting poor persistence of the SARS-CoV-2 gene fragments in wastewater.

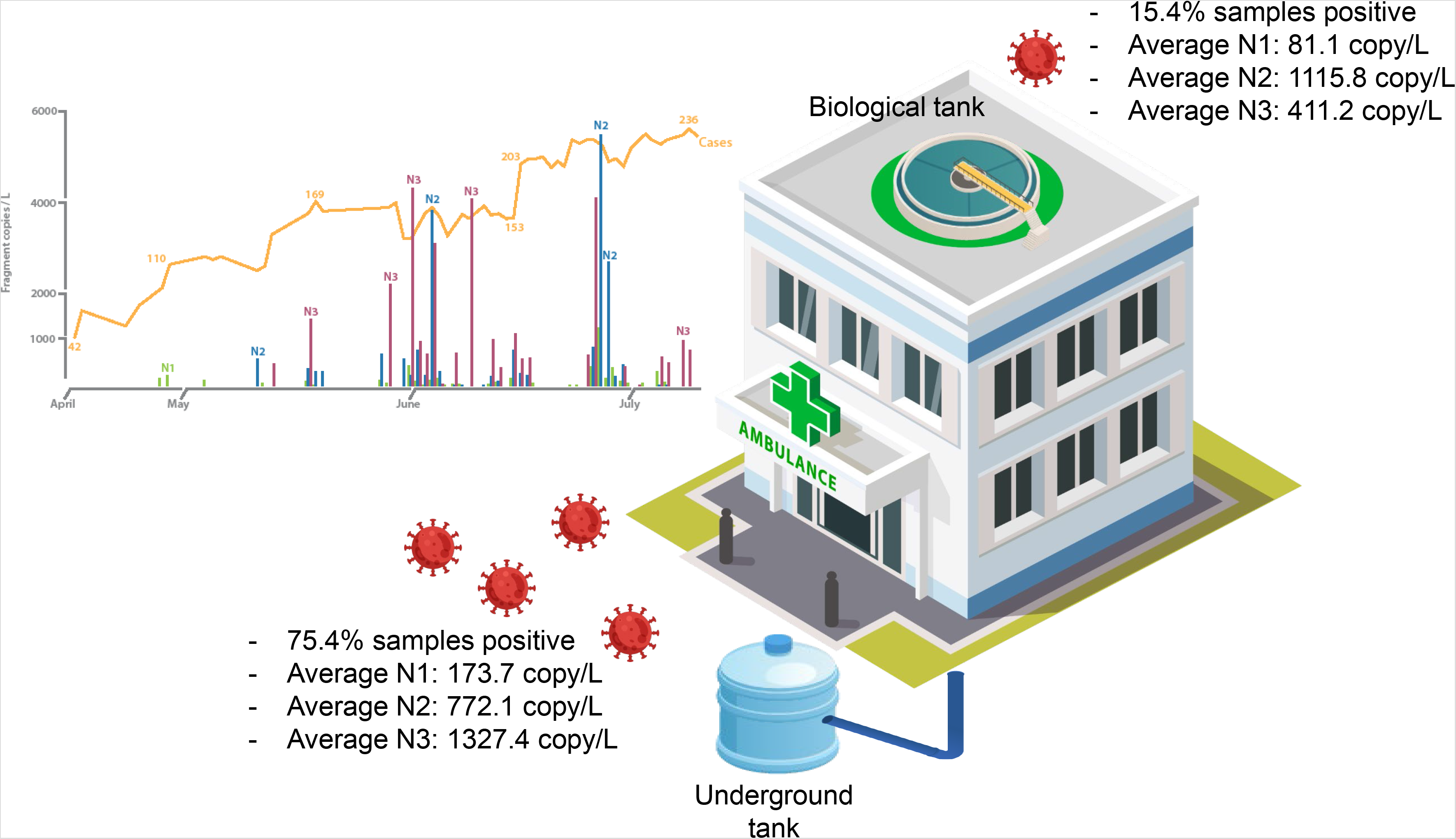

## Introduction

Data from the World Health Organization suggests that approximately 80% of Covid-19 infected individuals exhibit only mild symptoms or are asymptomatic^1^. In particular, asymptomatic cases are concerning as these individuals cannot be detected easily by the current swab testing protocols and can unknowingly transmit the virus to others. Although lockdown and curfew are very effective in slowing the spread of SARS-CoV-2, such intervention measures in the long term is unsustainable, and remains as a key but not the only solution to mitigating Covid-19. The other solution lies in increasing our testing capacity. However, to swab everyone in a country for clinical surveillance over a long term period is impractical considering the amount of resources and labor hours needed. In contrast, monitoring for SARS-CoV-2 directly in wastewaters over a longer duration would be an alternate method to complement clinical surveillance since about 39 to 65% of infected hosts, including asymptomatic carriers, shed the virus through their feces, while about 6% of patients shed it through urine^2, 3^.

By sampling for the waste stream generated from a community, we therefore have a composite sample that would be suitable for wastewater-based epidemiology (WBE). WBE is an approach that includes the qualitative and quantitative determination of biomarkers in raw wastewater to provide information on inhabitants within that wastewater catchment area. Monitoring for SARS-CoV-2 in wastewater has been demonstrated in many countries, including the Netherlands, Australia, Italy, Spain and US^4-9^. In these studies, SARS-CoV-2 was sporadically detected from the sampled wastewaters, and the reported abundance of SARS-CoV-2 in wastewater ranged from 19 to 2.2 × 10^8^ copies/L. In some instances, the WBE approach was able to detect SARS-CoV-2 in wastewater before clinically diagnosed cases were made known^10^. WBE hence demonstrates potential to serve as an early warning of re(emergence) of Covid-19 in communities^7^. However, without knowing the minimal number of positive cases needed in each community to achieve consistent detection of SARS-CoV-2 from the sewage networks, the definition of how early into the outbreak WBE can inform us remains rather unclear.

Ideally, the detection sensitivity of WBE can be defined by knowing the total number of symptomatic and asymptomatic individuals present in the community served by the wastewater treatment system. The volume of untreated sewage entering to the wastewater treatment plant (WWTP) should also be defined to estimate the total number of inhabitants served by the WWTP. While the latter requirement can be easily determined, the number of infected individuals is difficult to obtain unless an active surveillance is conducted to swab test all individuals, including asymptomatic ones, in the community. To circumvent this limitation, a controlled community, for example, hospitals providing treatment to Covid-19 patients can be used as a model. In this manner, the number of patients contributing SARS-CoV-2 to the hospital wastewaters are known through daily hospitalization and discharge records. Besides determining the detection sensitivity of WBE using hospital wastewaters as a study model, we further evaluate the correlation between detected abundance of SARS-CoV-2 gene fragments in wastewater with the number of patients. The abundance of nucleocapsid gene fragments as it moves from the underground septic tank to the first stage of the wastewater treatment plant situated on-site was also evaluated.

Findings from this study will provide inference to the minimal number of SARS-CoV-2 infected cases needed to generate quantitative PCR (qPCR) signals associated with the virus shed into a highly diluted wastewater stream. The correlation analysis provide insights to which of the nucleocapsid gene abundance would vary according to the number of infected individuals. Lastly, the persistence of the nucleocapsid genes in wastewater can be inferred by determining the changes in their abundance along different stages of the wastewater treatment plant.

## Materials and methods

### Quality control of sample processing and RNA extraction protocols

To determine recovery and viral nucleic acid extraction efficiency, a known concentration of murine norovirus (MNV) was spiked into untreated wastewater. MNV was spiked as surrogate because it is a positive-sense single-stranded RNA virus like SARS-CoV-2, and will likely be similar in terms of RNA extraction efficiency. Quantitative PCR (qPCR) for MNV was performed using forward primer (5’- CCGCAGGAACGCTCAGCAG-3’), reverse primer (5’- GGYTGAATGGGGACGGCCTG-3’) and Taqman probe (5’- FAM-ATGAGTGATGGCGCA-ZEN/IBFQ-3’) (Integrated DNA Technologies, Leuven, Belgium^11^. A six-points standard curve was generated using a synthetic oligonucleotide (gblocks^@^ gene fragment, Integrated DNA technologies, IA, USA) containing 300–350 bp DNA sequence that encompasses a complementary region for which the primers and probes would anneal to. An amplification efficiency of 102.7% was achieved. LOD for MNV qPCR reaction was 3.89-log copies/L and 10 copy/well, respectively. MNV is not anticipated to be present in municipal wastewaters and hence abundances detected by means of qPCR would be due to the spiking event.

### Sampling of hospital wastewaters from Jeddah, Saudi Arabia

Wastewater discharged from the hospital was collected at two sampling points of the wastewater treatment plant (WWTP) located on-site. The first sampling point was the underground septic tank and the second sampling point was from the aerobic biological tank located on the roof of the hospital. The daily wastewater intake for the on-site WWTP is 750 m^3^. The volume of wastewater generated by this hospital approximates the size of a community with ca. 2884 persons based on an average capita water usage of 260 L/person/d^12^. The hospital wastewater is treated first by biological activated sludge system, operated with hydraulic retention time of 9 h. The secondary-treated wastewater is then disinfected with 100 mg/L chlorine at a contact time of 2 h. Approximately 1 L of grab sample was individually collected from the underground tank (i.e., representing untreated wastewater) and from the top aqueous layer of the biological activated sludge tank (i.e., partially treated wastewater). The entire sampling duration was from 15 April to 9 July 2020, with frequencies ranging from 3 to 5 daily samples per week (Table S1). The lower sampling frequency during the early stages of the outbreak was due to the movement restriction order imposed in the city. Samples were stored at 4°C for not more than 1 week before they were processed.

### Processing of wastewater samples

250 to 500 mL of samples were individually concentrated for viral particles using the Millipore HA membrane method with slight modifications^13^. Briefly, 2 mL of 2.5 M MgCl2 was added to every 100 mL of the sample, agitated for 3 minutes and left to stand for 3 minutes. The samples were then filtered through the HA membrane. The HA membrane was then filtered through with 200 mL of 0.5 mM H2SO4 to adjust the isoelectric point of viruses. The viral particles retained on the HA membrane were eluted with 10 mL of 1 mM NaOH into a sterile collection tube that contained 100 µL of 100X tris-EDTA buffer and 50 µL of 100 mM H2SO4. The eluate was concentrated with Centripep YM-50 (Millipore) to approximately 680 µL, and an aliquot of 140 µL was extracted for its RNA using QIAmp Viral RNA kit (ThermoFisher Scientific, Carlsbard, CA). QIAmp Viral RNA kit is suggested by the US CDC as one of the extraction protocols to generate highly purified RNA^14^. Extracted RNA was converted to complementary DNA (cDNA) using Superscript III First Strand Synthesis System (ThermoFisher Scientific).

### Quantitative PCR

Genes associated with nucleocapsid (N) proteins N1, N2 and N3 of SARS-CoV-2 were targeted for quantitative PCR^15^. G-blocks that include the annealing regions of N1, N2 and N3 primers-probes were synthesized based on the published sequences of SARS-CoV-2^16^. Six-point standard curves were generated for each of the primer-probe pair to determine their respective amplification efficiencies. Limit of detection is estimated based on the lowest copy number of G-block template with detectable threshold Cq value. Based on the standard curves, the average amplification efficiency were 92.5%, 91.5%, 90.8% and the R^2^ value ranged from 0.98–0.99, 0.97–0.99, and 0.97–0.99 for N1, N2, and N3, respectively. The LOD was 5 copy/well, corresponding to 3.85-, 4.25-, and 3.35-log copies/L for N1, N2, and N3 qPCR reaction, respectively. A non-template control serves as negative control. A positive control made up from cDNA derived from RNA of SARS-CoV-2-positive clinical swab specimens was also included for qPCR. Positive controls have average Cq value of 29 for N1, 30 to 31 for N2 and N3 genes. Thermal cycling conditions include 50 cycles of 95°C for denaturation (3 s) and 55°C for annealing and amplification (30 s). All samples, standards and controls were performed in technical duplicates.

## Results and discussion

### Recovery and extraction efficiency

A total of 6.89-log copies/L MNV was spiked into the untreated wastewater. The overall recovery and RNA extraction efficiency of murine norovirus was ca. 45%, and falls within the range reported by earlier studies (ranging from 10% to 73%) albeit using different concentration procedures. For example, direct ultracentrifugation of 20 mL of wastewater through a 50% sucrose phase layer recovered 12% of the deactivated SARS-CoV-2 spiked into the sample^17^. Aluminium hydroxide adsorption-precipitation (adjusted to pH 6) method recovered on average 11% of spiked porcine epidemic diarrhea virus and mengovirus, respectively, from 200 mL wastewater^8^. Ultrafiltration of 100–200 mL using Centricon Plus-70 (molecular weight cut-off of 10 kDa) recovered an average 73% of the spiked F-specific RNA phages but with high standard deviation of 50%^7^.

In addition, the reported differences in recovery efficiencies can be due to the varying type of viral surrogates spiked. Different viruses have different isoelectric points (pI), which are the pH values at which the net surface charge switches. The determination of pI is particularly relevant for concentration protocols that utilize electrocharged membranes or coagulant adsorption-precipitation. Norovirus was previously reported to have a pI ranging from 5.5 to 6^18^, while the spike protein of porcine epidemic diarrhea virus has a pI of 5.0^19^. In contrast, there is no experimental data currently available that reports on the pI of SARS-CoV-2 but predictive calculations for some of its key proteins put the range of pI between 4.2 to 10.1^20^. It is therefore uncertain which particular surrogate used thus far among studies would best approximate to SARS-CoV-2.

Regardless of the viral surrogates, recovery of viral particles from untreated wastewater has been challenging as evidenced from the wide range of reported efficiencies. High turbidity and organic carbon content in raw wastewater matrix can foul ultrafiltration membranes and decrease flux. In this study, the raw wastewater samples had an average dissolved organic carbon (DOC) concentration of 6.4 mg/L and average turbidity of 10.6 NTU (Table 1). DOC can facilitate unintended adsorption of virus onto the filtering glassware^21^, and hence lower the probability of recovering viral particles that are already present in low abundance and/or not homogenously distributed in the wastewater matrix. Furthermore, suspended particles that accounted for turbidity can clog the membranes rapidly and permit only a limited volume of untreated wastewater (< 500 mL) to be filtered through without significantly lengthening the processing time.

**Table 1.**
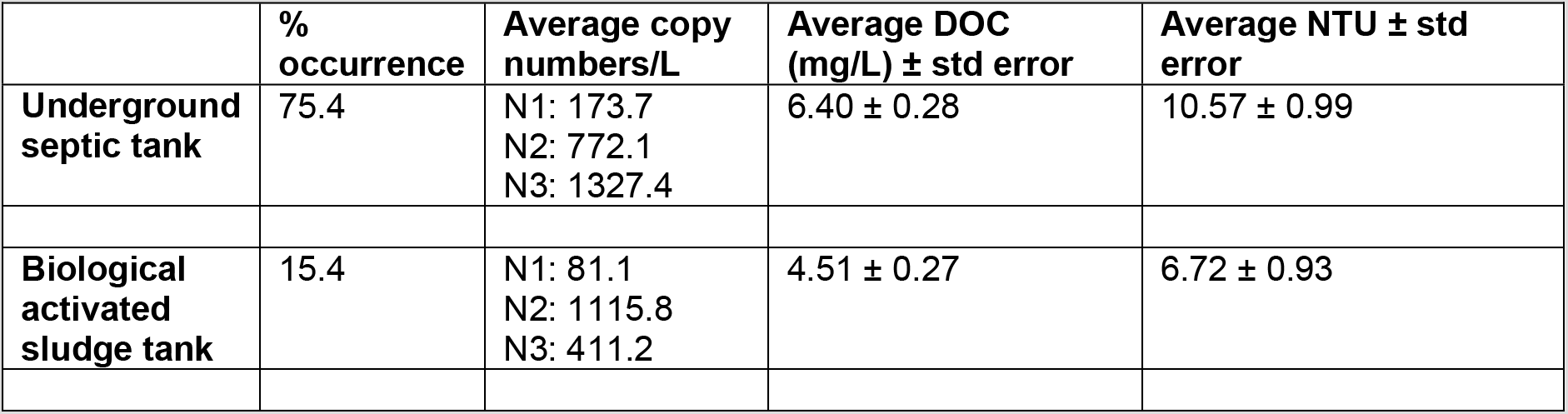
Summary of the percentage of samples detected positive for nucleocapsid genes of SARS-CoV-2 and the average abundance among positive detection. Average dissolved organic carbon and turbidity ± standard error were also measured for wastewaters collected at both sampling points.

| | %<br>occurrence | Average copy<br>numbers/L | Average DOC<br>(mg/L) ± std error | Average NTU ± std<br>error |
| --- | --- | --- | --- | --- |
| **Underground**<br>**septic tank** | 75.4 | N1: 173.7<br>N2: 772.1<br>N3: 1327.4 | 6.40 ± 0.28 | 10.57 ± 0.99 |
| **Biological**<br>**activated**<br>**sludge tank** | 15.4 | N1: 81.1<br>N2: 1115.8<br>N3: 411.2 | 4.51 ± 0.27 | 6.72 ± 0.93 |

### Detected abundance of SARS-CoV-2 in untreated hospital wastewaters

43 out of the 57 collected samples of untreated hospital wastewaters (i.e., 75.4%) tested positive for the nucleocapsid genes of SARS-CoV-2. Compared to earlier studies which reported occurrence frequency to range from 15.6 to 50% of the collected municipal wastewaters^4, 6, 9, 22, 23^, the number of samples that were positive for SARS-CoV-2 occurred at higher frequency in this study. This is not surprising since the wastewaters originated from a hospital that provides treatment for Covid-19 infected patients. However, despite the confirmed presence of infected patients in this hospital since our first sampling date, we did not detect SARS-CoV-2 in the untreated wastewaters sampled on 15, 16, 22 and 24 April 2020. Instead, a relatively consistent detection of the SARS-CoV-2 N1 gene was only observed after 27 April 2020. Wu et al. determined correlation between wastewater viral titers and daily reports of new clinically confirmed Covid-19 cases in Boston, US, and found that the best correlation was seen when comparing new clinical cases back-dated by 4 days^24^. Hence, it is likely that a consistent detection of SARS-CoV-2 in the hospital wastewaters can be achieved when the cumulative number of Covid-19 patients in the hospital was > 73 (Figure 1). Considering that the daily wastewater intake for the on-site hospital WWTP is 750 m^3^, the volume of wastewater generated by this hospital approximates the size of a community with ca. 2884 persons based on an average capita water usage of 260 L/person/d^12^. This therefore suggests that the procedure described in this study has a detection sensitivity of ca. > 253 infected persons per 10,000 inhabitants.

**Figure 1.**
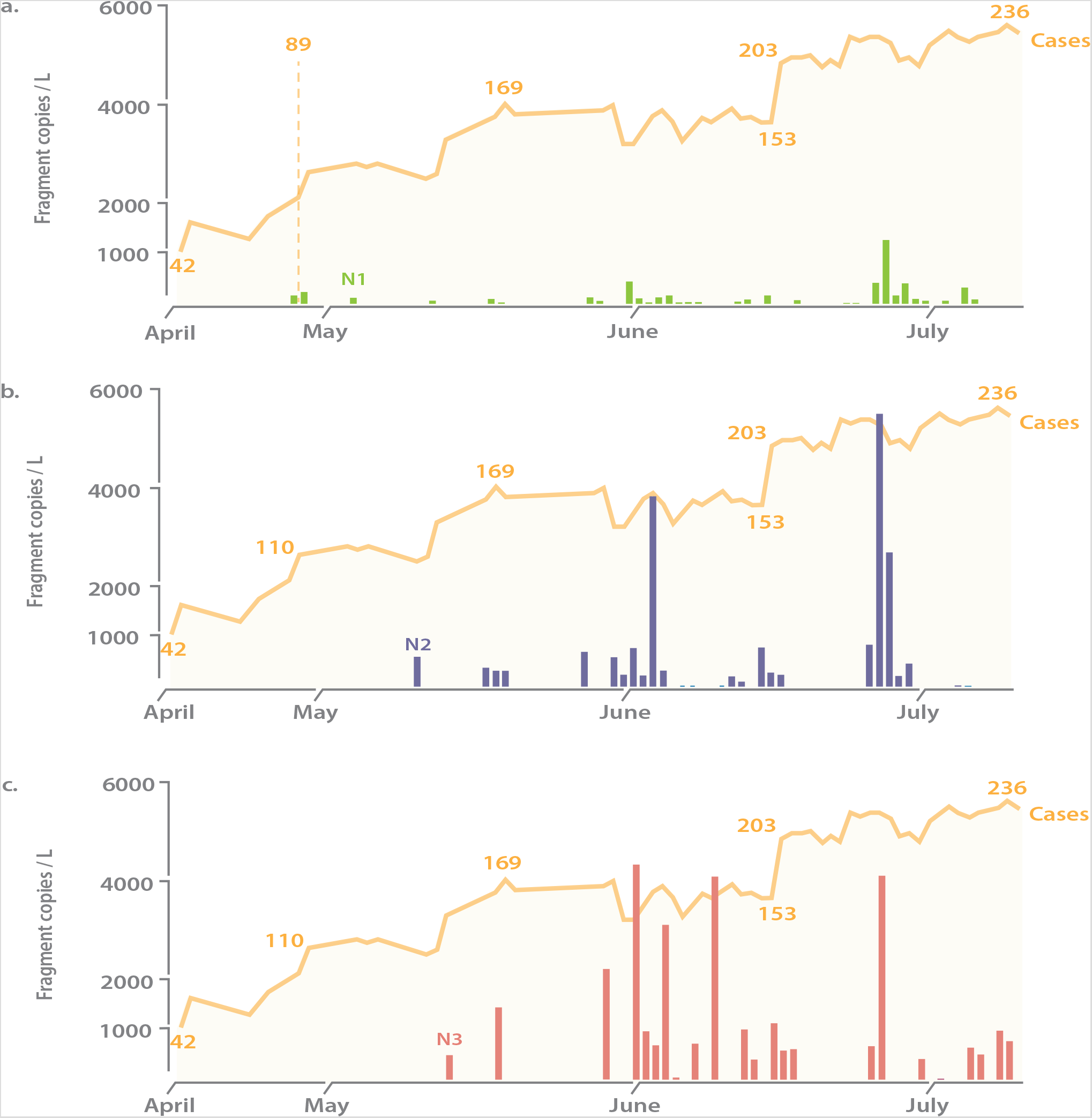
Abundance of nucleocapsid genes associated with SARS-CoV-2 and the number of hospitalized patients throughout the sampling period. (a) N1 gene, (b) N2 gene, (c) N3 gene. The dashed line in panel (a) indicates the number of hospitalized patients needed to obtain qPCR signals from this wastewater matrix.

Similar to that reported by Medema et al. (2020), N1 and N2 genes were observed in the hospital wastewater first before N3 gene. The simultaneous detection of N1, N2 and N3 gene was observed after a cumulative 138 patients or more were hospitalized (i.e., > 464 infected persons per 10,000 inhabitants). With the exception of N1 and N2 gene exhibiting a weak correlation (r = 0.21 and 0.24, respectively), there was no apparent correlation between abundance of N3 genes and the number of hospitalized patients. The weak correlation and sporadic detection of SARS-CoV-2 genes in hospital wastewaters may be dependent on the viral shedding load per patient and the shedding duration. In this study, information on whether patients are exhibiting gastrointestinal distress was not provided to us due to concerns of breaching patients’ confidentiality. Hence it is uncertain how many percentage of the patients may be shedding the virus into the wastewater to result in the sporadic detection of SARS-CoV-2.

### Log reduction of SARS-CoV-2 gene fragments at different stage of treatment process

The biological activated sludge tank is an open-air tank placed on the roof of the hospital, and wastewater inside the tank was subjected to an increasing ambient temperature and solar irradiance during the sampling period as KSA enters into summer. To exemplify, maximum diurnal temperature increased from 41.5°C in April to 45.3°C in May and 46.3°C in June 2020, while minimum diurnal temperature also increased from 14.2°C in April to 21°C in June (personal communication, Matthew McCabe). The maximum net solar irradiation measured at 1 pm also increased from 1184.9 W/m^2^ in April to 1333.0 W/m^2^ in June (personal communication, Matthew McCabe). Parameters related to water quality, namely DOC and turbidity were lower in values for wastewater sampled from the biological activated sludge tank compared to the underground septic tank (Table 1). This indicates a certain degree of partial treatment was achieved at the sampling point of the activated sludge tank.

Likewise, this level of partial treatment resulted in a lower detection frequency of SARS-CoV-2 in wastewaters sampled from the biological activated sludge tank. Only 8 out of 52 samples (15.4%) collected from the biological activated sludge tank were tested positive for N genes associated with SARS-CoV-2. Specifically, N1 genes were detected in 5 samples, N2 genes were detected in 4 samples, and 1 positive occurrence of N3 genes among the collected samples. N1 genes were detected first before N2 and N3. The average abundance of N1 and N3 genes decreased by 0.3-log (i.e., 50%) and 0.5-log(i.e., 70%), respectively, from the underground septic tanks to the biological activated sludge tank. However, there was an increase by 44% in the average abundance of N2 genes sampled from the activated sludge tank compared to that in the underground septic tank. This suggest a potential accumulation of N2 genes in the activated sludge tank, likely due to better persistence of N2 compared to N1 and N3 genes. Most of the positive detection in the biological activated sludge tank occurred in late June, and coincides after a period of high cumulative numbers of hospitalized patients. Despite the slight increase of N2 genes in a small group of samples, it is unlikely that the N genes will persist in the final treated wastewater since the last stage of the hospital WWTP includes a final disinfection step at a very high chlorine concentration (100 mg/L) and at long contact time (2 h). An earlier study determined that 6.5 mg/L of free chlorine and 1.5 h of contact time was already sufficient to remove 0.5 to 18.7 × 10^3^ copies/L of SARS-CoV-2 viral RNA to levels below detection limits^25^.

## Conclusion

By utilizing a confined environment (i.e., a hospital with known number of Covid-19 cases), this study determined the detection sensitivity of wastewater-based epidemiology for SARS-CoV-2 to be about > 253 persons per 10,000 inhabitants. The correlation between nucleocapsid gene abundances of SARS-CoV-2 and the number of infected hospitalized patients was weak, suggesting the difficulty in correlating the number of cases based on the gene abundances detected in wastewaters. However, it is likely that when the nucleocapsid genes were detected, there is already a substantial number of cases circulating in the community that would warrant immediate intervention measures to be taken. N genes do not persist well in the hospital wastewater based on the lower detection frequency in partially treated wastewater. In instances where the nucleocapsid genes were detected in the partially treated wastewater, N1 and N3 genes had already decreased by 0.3-log and 0.5-log (i.e., ca. 50 and 70%, respectively). This is with the exception of N2 genes that may persist better than N1 and N3. Future studies would be needed to improve the recovery efficiency of viral particles from wastewater matrix, and to determine the decay kinetics of the N genes in various environmental conditions to facilitate the detection sensitivity and capabilities of WBE.

## Data Availability

All data referred to in the manuscript are included and additional data can be requested from corresponding author.

## Acknowledgements

This work was funded by KAUST baseline grant BAS/1/1033-01-01 awarded to PYH. The authors would like to thank the medical team of the studied hospital for providing access to wastewater samples and for saving lives during this pandemic. The authors would also like to thank Professor Matthew McCabe and Mr Samir Almashharawi for providing the ambient temperature and solar irradiance measured at the Hada Al Sham monitoring station.

## Supporting materials available

Table: Information on the samples collected for this study.

## TOC

**Table S1**. Information on the sampling dates at the respective sampling point……………………………..1

**Table S1.**
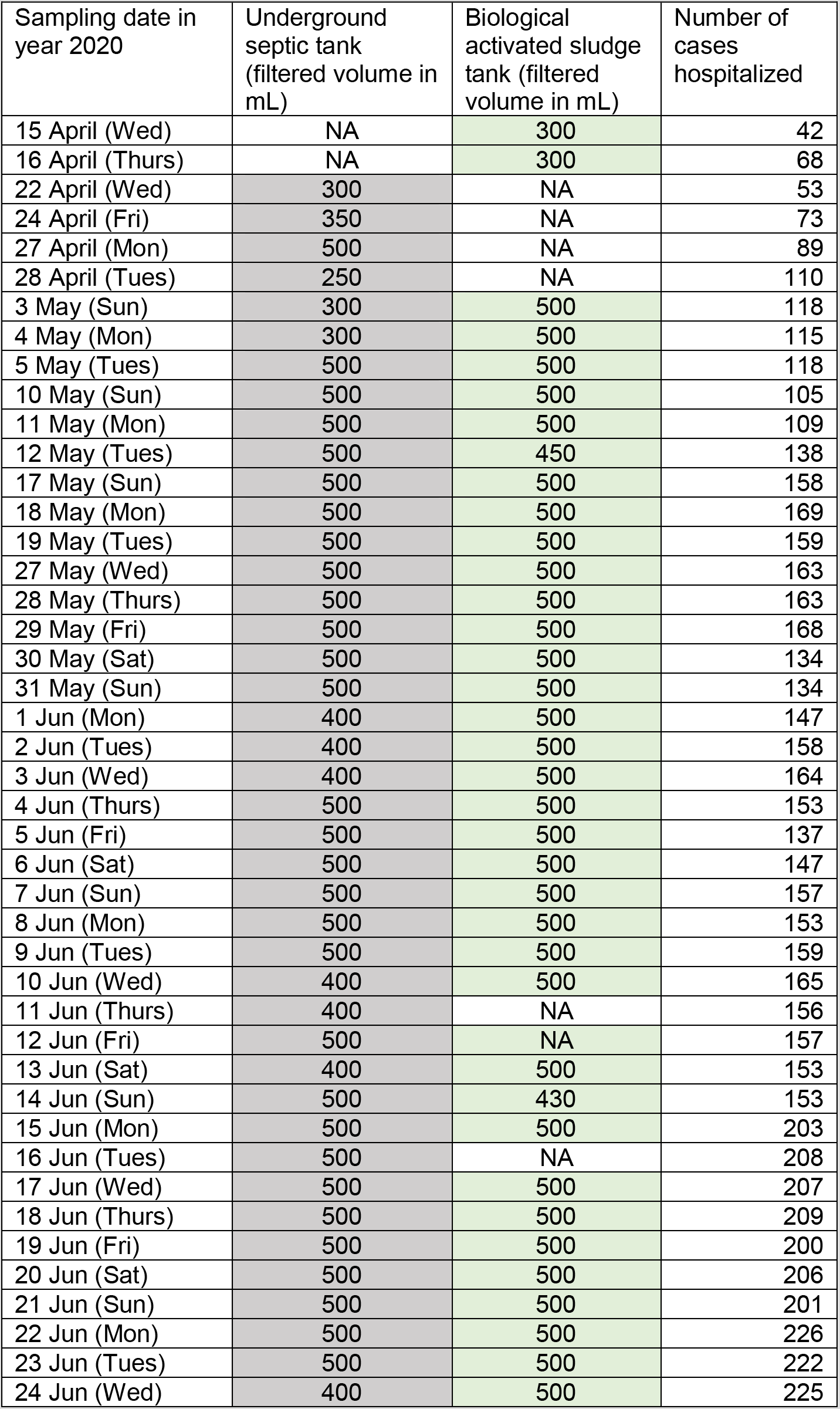
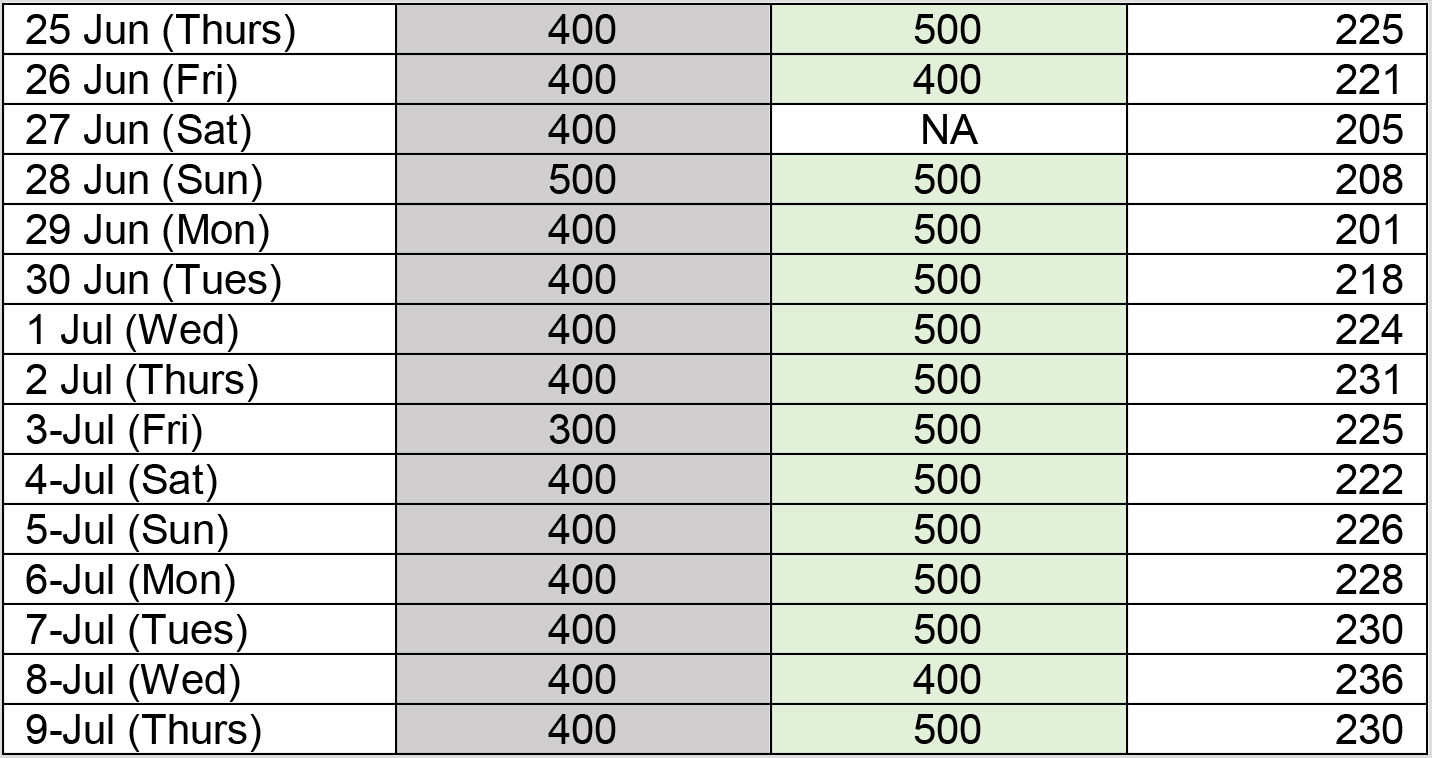
Information on the sampling dates at the respective sampling point. Gray cell indicates sampling was performed at the septic tank on that particular date. Green cell indicates sampling was performed at the biological tank on that particular date. Values in each cell refer to the filtered volume.

| Sampling date in year 2020 | Underground septic tank (filtered volume in mL) | Biological activated sludge tank (filtered volume in mL) | Number of cases hospitalized |
| --- | --- | --- | --- |
| 15 April (Wed) | NA | 300 | 42 |
| 16 April (Thurs) | NA | 300 | 68 |
| 22 April (Wed) | 300 | NA | 53 |
| 24 April (Fri) | 350 | NA | 73 |
| 27 April (Mon) | 500 | NA | 89 |
| 28 April (Tues) | 250 | NA | 110 |
| 3 May (Sun) | 300 | 500 | 118 |
| 4 May (Mon) | 300 | 500 | 115 |
| 5 May (Tues) | 500 | 500 | 118 |
| 10 May (Sun) | 500 | 500 | 105 |
| 11 May (Mon) | 500 | 500 | 109 |
| 12 May (Tues) | 500 | 450 | 138 |
| 17 May (Sun) | 500 | 500 | 158 |
| 18 May (Mon) | 500 | 500 | 169 |
| 19 May (Tues) | 500 | 500 | 159 |
| 27 May (Wed) | 500 | 500 | 163 |
| 28 May (Thurs) | 500 | 500 | 163 |
| 29 May (Fri) | 500 | 500 | 168 |
| 30 May (Sat) | 500 | 500 | 134 |
| 31 May (Sun) | 500 | 500 | 134 |
| 1 Jun (Mon) | 400 | 500 | 147 |
| 2 Jun (Tues) | 400 | 500 | 158 |
| 3 Jun (Wed) | 400 | 500 | 164 |
| 4 Jun (Thurs) | 500 | 500 | 153 |
| 5 Jun (Fri) | 500 | 500 | 137 |
| 6 Jun (Sat) | 500 | 500 | 147 |
| 7 Jun (Sun) | 500 | 500 | 157 |
| 8 Jun (Mon) | 500 | 500 | 153 |
| 9 Jun (Tues) | 500 | 500 | 159 |
| 10 Jun (Wed) | 400 | 500 | 165 |
| 11 Jun (Thurs) | 400 | NA | 156 |
| 12 Jun (Fri) | 500 | NA | 157 |
| 13 Jun (Sat) | 400 | 500 | 153 |
| 14 Jun (Sun) | 500 | 430 | 153 |
| 15 Jun (Mon) | 500 | 500 | 203 |
| 16 Jun (Tues) | 500 | NA | 208 |
| 17 Jun (Wed) | 500 | 500 | 207 |
| 18 Jun (Thurs) | 500 | 500 | 209 |
| 19 Jun (Fri) | 500 | 500 | 200 |
| 20 Jun (Sat) | 500 | 500 | 206 |
| 21 Jun (Sun) | 500 | 500 | 201 |
| 22 Jun (Mon) | 500 | 500 | 226 |
| 23 Jun (Tues) | 500 | 500 | 222 |
| 24 Jun (Wed) | 400 | 500 | 225 |
| 25 Jun (Thurs) | 400 | 500 | 225 |
| 26 Jun (Fri) | 400 | 400 | 221 |
| 27 Jun (Sat) | 400 | NA | 205 |
| 28 Jun (Sun) | 500 | 500 | 208 |
| 29 Jun (Mon) | 400 | 500 | 201 |
| 30 Jun (Tues) | 400 | 500 | 218 |
| 1 Jul (Wed) | 400 | 500 | 224 |
| 2 Jul (Thurs) | 400 | 500 | 231 |
| 3-Jul (Fri) | 300 | 500 | 225 |
| 4-Jul (Sat) | 400 | 500 | 222 |
| 5-Jul (Sun) | 400 | 500 | 226 |
| 6-Jul (Mon) | 400 | 500 | 228 |
| 7-Jul (Tues) | 400 | 500 | 230 |
| 8-Jul (Wed) | 400 | 400 | 236 |
| 9-Jul (Thurs) | 400 | 500 | 230 |

## Notes

### Competing Interest Statement

The authors have declared no competing interest.

### Clinical Trial

The study only sampled wastewater and does not involve any clinical trials.

### Funding Statement

This study is funded by KAUST Baseline funding BAS/1/1033-01-01 awarded to Peiying Hong

### Author Declarations

King Abdullah University of Science and Technology Institutional Biosafety and Bioethics Committtee has provide approval for the research described involving wastewater sampling and monitoring.

